# The Impact of the Performance-Based Financing Project: An Observational Panel Study on Health Workers’ Output in Rural Mezam, Northwest Region, Cameroon

**DOI:** 10.1101/2024.11.20.24317631

**Authors:** Therence Nwana Dingana, Balgah Roland Azibo, Daniel Agwenig Ndisang, Stewart Ndutard Ngasa, Leo Fosso Fozeu

## Abstract

Performance-Based Financing (PBF) has been implemented in many countries to improve healthcare access, quality, and outcomes while ensuring the efficient and equitable allocation of resources within the healthcare system. However, very little effort is visible in assessing its real impacts. This study evaluates the impact of the PBF project on health workers’ output and healthcare quality in the Mezam division in the North West region of Cameroon between 2012 and 2022. Specifically, the study aims to understand health workers’ perceptions of the PBF project, analyze the effect of PBF on health workers’ output, and examine the impact of PBF on healthcare quality. A structured questionnaire was used to generate panel data among healthcare workers in six beneficiary health districts in the study site. The perception scores were estimated based on the Net Promoter Score (NPS) methodology and variability tested using ANOVA. Health workers’ output and performance indicators were analyzed using the chi-square test, assessing the relation between PBF introduction and changes in health workers’ output, while healthcare quality metrics were analyzed using the Mann-Whitney U test to compare healthcare quality before and after PBF implementation. The results showed that health workers’ perceptions varied but were generally positive, with a Net Promoter Score (NPS) of approximately 48.25. PBF significantly boosted health workers’ output (p = 0.002) and healthcare quality (p < 0.05). It can be concluded that the PBF project in the Mezam division had positive effects on workers’ output and healthcare quality. Given the positive impacts, the study recommends scaling up PBF initiatives in Cameroon and other African countries with precarious health systems. Our study demonstrates the relevance of impact assessments in providing evidence for making informed decisions on efficient resource allocation in the health sector.

## INTRODUCTION

Access to health is central to human survival. Its emphasis among the Sustainable Development Goals (SDGs), particularly SDG 3, which aims to ensure healthy lives and promote well-being for all ages is justified. Achieving this goal necessitates significant investment in health initiatives, especially in developing countries where health systems are largely inadequate (1). Low- and middle-income countries (LMICs) face increasing healthcare demands but have limited financial resources, allocating only 5-7% of GDP to healthcare compared to 12.5% in high-income countries (2). Sub-Saharan African (SSA) countries spend even less: not more than 4-5% of the GDP (3). Universal Health Coverage (UHC) is a key strategy to optimize health sector expenditure and achieve SDG 3 by 2030 (4), particularly in SSA, where over 408 Million of the 600 Million people in Africa who lack access to basic health care services are lodged (5); (6). This involves cost-effective, context-specific strategies such as health financing, health insurance, prepayment schemes, and innovative provider payment mechanisms (7); (8); and (9).

Performance-Based Financing (PBF), also known as Result-Based Financing or Pay for Production, is a health financing scheme designed to increase the quality and quantity of healthcare services by linking payments in health facilities based on performance (10); (11). Performance-Based Financing (PBF) is a health financing approach used to improve the quantity and quality of healthcare services in low- and middle-income countries (LMICs) by linking financial rewards to predefined targets (12). This approach incentivizes healthcare providers, thereby promoting better performance (7). PBF enhances the autonomy of healthcare facilities and involves a comprehensive performance framework, including regulatory functions, contract development and verification agencies, and community empowerment initiatives (13) (14). It aims to contain costs and ensure sustainable revenue through a mix of cost recovery, government contributions, and international support (15)

In Cameroon, PBF targets health facilities and workers, aiming to improve healthcare service utilization and quality at various system levels (16) (17). Research has shown that health workers receiving PBF payments reported feeling motivated by performance bonuses, leading to improvements in working environments, including enhanced supervision and availability of essential resources (18). Both financial and non-financial performance-based incentives have been found to boost health worker motivation and performance in various settings (19).However, the actual impact of PBF schemes on achieving better healthcare quality outcomes remains uncertain (20) (21). Supported by major international donors such as the WHO, World Bank, and USAID, PBF programs have shown potential in improving service coverage and quality in various LMICs, despite some challenges like resource allocation and evaluation issues (22) (23). Evaluations in countries like Rwanda, Argentina, Cambodia, Tanzania, and Zambia have highlighted PBF’s potential to enhance service coverage and quality (24) (25) (26).

This study examines the effects of PBF on health workers’ output in Mezam division, Northwest Region of Cameroon, an area with unique healthcare challenges, including inadequate infrastructure, insufficient personnel, and disparities in urban-rural healthcare access (4). Healthcare workers in Mezam operate in resource-constrained environments with limited training, equipment, and motivation. Ensuring their motivation and high-quality care delivery is crucial to improving health outcomes in the region.

The global focus on health challenges in LMICs has intensified with the onset of the sustainable development goals on universal health and well-being toward 2030 (27). Strengthening health systems in these regions like SSA, is crucial but complicated by limited resources (28). Cameroon faces a low health worker-to-patient ratio, with approximately 0.4 physicians and 1.6 nurses per 1,000 people, which is below the regional average (1). Dissatisfaction with wages has led to abandonment of duties, strikes; necessitating the government and World Bank to introduce the PBF program in 2011 to improve healthcare delivery (29). Despite its scale-up to national policy by 2019, the program’s impact in the conflict-affected Northwest region remains under-evaluated. This study aims assesses the effects of PBF on health workers’ output in Mezam division in the North West region of Cameroon as a first step in reducing this existing research gap. Specifically, the study seeks to understand health workers’ perception of the PBF project, analyze the effects of PBF on health workers’ output and assess the impact of PBF on the quality of healthcare in the study area.

Three hypotheses guide the study as follows:

H1 There is a significant improvement in health workers’ perception of the PBF project.
H2 PBF has a positive impact on health workers’ output in beneficiary health centers.
H3 There is a significant difference in the quality of healthcare services before and after implementing PBF in the study site.

The contribution of this study is twofold. First, it adds to the slow-growing empirical studies on the effects of PBF on the health sector in SSA. By examining how PBF influences healthcare workers’ productivity and care quality, providing critical data for quality improvement and evidence-based decision-making. By examining the intricate interplay between PBF and health worker performance in Mezam, this study aspires to contribute to the achievement of Universal Health Coverage (UHC) and the Sustainable Development Goals (SDGs) (15). The findings will guide policymakers, healthcare administrators, and funding agencies in optimizing health investments and financing strategies. Second, the study reiterates the relevance and nuances of undertaking impact assessment of health care projects.

This article proceeds as follows. The next section reviews the literature on health workers’ perceptions of PBF. As will be observed, health workers generally have positive perceptions of PBF projects. This is followed by an overview of the topical literature on the effect of PBF on health workers’ output and quality of care. PBF increases productivity and health service utilization and drives health workers’ performance compared to the situation in its absence.

### Health workers’ perception of Performance Based Financing

Health workers in beneficiary health units perceive the PBF project positively but with nuanced views shaped by various factors. Bhatnagar & George found out that PBF payments enhanced their working environments, including better supervision and availability of essential drugs, creating a positive influence (18). The need to understand the complex motivational mechanisms at play has been highlighted, as PBF impacts health workers’ attitudes and behaviors. In Zambia, PBF improved job satisfaction and retention rates, indicating a favorable reception (30). Gergen reported that improved resources and management support in Mozambique fostered self-efficacy and pride among health workers (31). It has been emphasized that leadership and organizational capacity in Burkina Faso significantly influenced health workers’ motivational reactions (10). Overall, PBF is generally perceived positively, enhancing motivation and work conditions, though its effectiveness is contingent on supportive organizational contexts.

### Effect of Performance based financing on health workers’ output and quality of care

PBF positively impacts health workers’ output by providing financial incentives, strengthening management, and enhancing clinical capacities through regular supervisions (21). In Benin, a study found that PBF motivates health workers and fosters a culture of continuous improvement, leading to better performance (32). Similarly, PBF has been shown to increase productivity and health service utilization by encouraging health workers to meet performance targets (33). In Tanzania, training programs associated with PBF have been observed to improve the quality of healthcare delivery. Studies conducted in Burkina Faso have reiterated that PBF incentivizes health workers to meet specific goals, thereby enhancing their output and the quality of care (34). Additionally, research in Zambia emphasized the importance of understanding the mechanisms through which PBF influences satisfaction, motivation, and retention of health workers (30). Overall, PBF enhances health workers’ performance by aligning incentives with performance targets and investing in supportive mechanisms such as training and supervision.

## MATERIALS AND METHODS

### Background of Study Area

This study was carried out in Mezam, the central and most populated division in the Northwest region of Cameroon. It has 654,659 inhabitants (35), 62 health areas, and 1,480 health workers (36). The region is currently emerged in a socio-political crisis that turned violent in November 2016, causing significant displacement and affecting livelihoods (37), including access to healthcare services. The healthcare workforce ratios in Mezam division are below WHO recommendations, with 1 doctor per 4,340 people and 1 nurse per 1,592 people (38). Access to healthcare facilities varies, with some residents traveling long distances for services (17). Health insurance coverage remains limited, with many relying on out-of-pocket payments (39). The healthcare infrastructure includes a mix of public and private facilities, with urban areas generally better equipped. Mezam faces various health challenges, including infectious diseases and maternal and child health issues. The scaling up of the PBF project in 2018 was anticipated to boost the collapsing health system (4).

### Study Design

This quantitative panel study involved healthcare providers (nurses, midwives, doctors, laboratory technicians) who worked before and during the PBF implementation. The panel study method was chosen to examine changes in healthcare workers’ perceptions, output, and healthcare quality over time. A total of 315 health workers in 20 health facilities were selected using a multi-stage stratified random sampling approach.

### Population and Sampling

The study population included healthcare workers from Mezam Division who had worked both before and during the PBF implementation. Those with PBF experience outside the Northwest region or who worked only during the PBF period were excluded. Workers transferred within Mezam Division with PBF experience were included, ensuring a comprehensive understanding of both periods. A multistage sampling design was used to ensure representativeness in recruiting participants. First, Mezam Division was purposefully chosen due to its relevance as the PBF project’s focus area. Sixteen public, private, and confessional health centers actively engaged in the PBF project were then purposively selected to ensure diversity. Next, they were categorized into rural and urban strata and sampled proportionally. Health workers were selected based on stratified sampling to ensure specialty representation. In centers with more than five specialties, at least four were selected, with at most five participants per specialty included, with a cap of 20 total participants per facility. The sample size was determined using Taro Yamane’s formula for finite populations n = N/{(1+N (e)^2^}. With a total of 1,480 healthcare workers and a significance level of 0.05, the sample size was calculated to be 315 health workers. A proportionate fraction per health district based on the total number of health workers was used for participant recruitment.

### Method and Source of Data Collection

Data were collected between mid-May and mid-July 2023, using a structured questionnaire, pre-tested for clarity and reliability. The questionnaire covered identification details of healthcare workers and facilities, socio-demographic characteristics (age, gender, education, and experience), perceptions of the PBF project, its effects on healthcare workers’ output, and its impact on healthcare quality. Face-to-face interviews were conducted by trained field staff, ensuring consistent questionnaire administration. Interviews were conducted privately to promote honest responses, and all answers were recorded for analysis. This method allowed the collection of comprehensive data on the PBF project’s influence on performance and healthcare delivery. Both primary data (questionnaire responses) and secondary data (books, reports, journal articles) were used. Primary data provided specific insights, while secondary data offered a broader theoretical context.

### Validity and reliability

Content validity was ensured through expert review and a pilot study to refine the questionnaire. The instrument was examined and modified to achieve 98% validity.

Reliability was tested through a pilot study with five questionnaires administered to Tubah district hospital staff. The analysis confirmed the questionnaire’s reliability.

### Data Analysis

To evaluate perceptions, mean perception scores were calculated, and the net perception score was used for the final analysis. The significance of its variability by socio-demographic characteristics was tested using the ANOVA test. The Chi-square test assessed the association between PBF and health workers’ output. The Mann-Whitney U test compared healthcare quality before and after PBF implementation.

### Ethical Consideration

Ethical clearance was obtained from the Regional Ethical Committee for Human Health Research Northwest Region. Administrative clearance and verbal consent from health facility heads were also secured. Participants were informed about the study’s aim and their voluntary participation. Confidentiality was ensured, and data were stored in a password-protected database accessible only to the primary researcher and supervisor.

## RESULTS AND DISCUSSION

### Socio-demographic characteristics

The study sampled 315 health workers in the Mezam division. A face-to-face questionnaire was administered. Descriptive data collected was then analyzed and is presented thus; The study sample of 315 healthcare workers was mainly from Bamenda (41.9%), followed by Tubah (16.2%) and Santa (15.2%). The majority were aged 30–39 years (37.8%), with 25.1% under 30. Most participants were female (74.6%). Educationally, 63.2% had university degrees, 19.4% held post-graduate qualifications, 14.0% had secondary education, and 3.5% had primary education. Professionally, nurses comprised 61.3% of the sample, while laboratory technicians (8.3%), midwives (11.4%), and medical doctors (2.5%) were less common. Other roles included biomedical technicians/imaging (1.0%) and auxiliary staff (15.6%).

### Healthcare Worker Characteristics Before and with Performance Based Financing

The study examined health practitioners’ employment status and other demographic factors before and after the introduction of the PBF program (Table 1). Government employment increased from 40.3% to 45.4%, while private sector employment dropped from 48.9% to 40%. Employment in confessional sectors rose from 10.8% to 14.6%. Health centers saw a decrease in workforce, from 24.1% to 18.1%, and MHCs from 17.5% to 12.4%, while district hospitals and regional hospitals saw increases of 4.8% and 7.3%, respectively. Christian respondents slightly declined, while Muslim representation increased. Marital status shifts included a rise in married respondents from 56.5% to 61% and a decrease in singles from 39.4% to 34.6%. Longevity in years of service saw a shift from a median of 5 years pre-PBF to 9 years post-PBF, with increased experience among respondents. Monthly incomes also rose, with median income increasing from 60,000fcfa to 75,000fcfa post-PBF. Practitioners earning less than 50,000 FCFA dropped from 23.2% to 13%, while those earning over 200,000 FCFA rose from 1.3% to 4.8%. Household sizes largely remained the same, but the number of households with more than six persons increased.

**Table 1:**
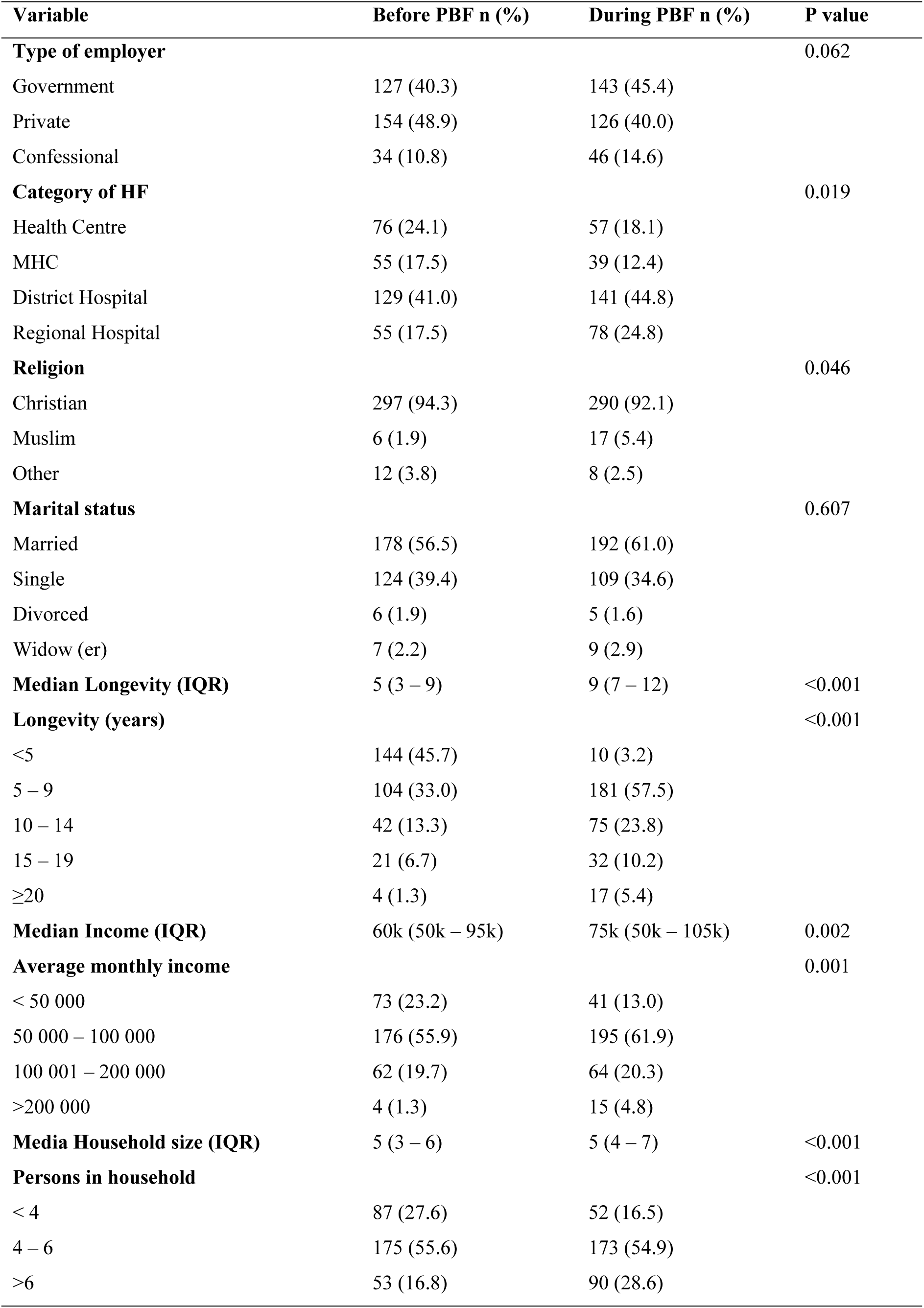
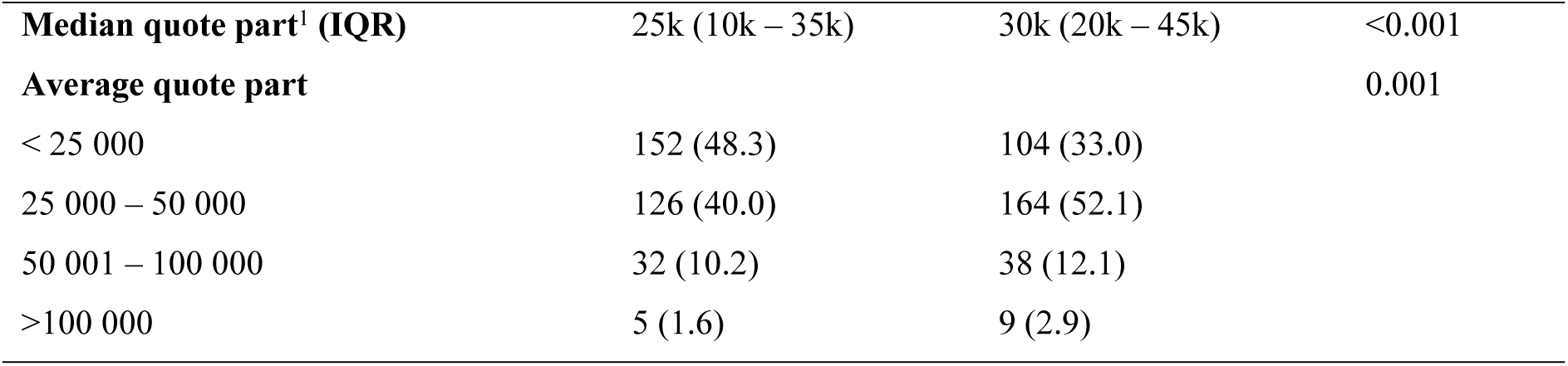
Presentation of results related to variables in the study.

The median "quote part" income rose from 25,000fcfa to 30,000fcfa. The term "quote part" in the context of health workers typically refers to a portion or share of something, often financial incentives or rewards, that is allocated to individual health workers based on certain criteria or performance indicators. In the context of Performance-Based Financing (PBF) or similar incentive programs, a "quote part" for health workers would represent the portion of the financial incentives or bonuses that each health worker is entitled to receive based on their performance in achieving predefined targets and objectives.

### Healthcare workers’ perceptions and attitudes toward the Performance Based Financing project

An assessment of practitioners’ perceptions of the PBF revealed diverse opinions as presented in Table 2Perception of Performance Based Financing

**Table 2:** Perception of Performance Based Financing.

| **Variables** | **Number** | **Percentage** |
| --- | --- | --- |
| **How well have your expectations from PBF been met** |  |  |
| Very unsatisfied | 27 | 8.6 |
| Unsatisfied | 76 | 24.1 |
| Neutral | 105 | 33.3 |
| Satisfied | 82 | 26.0 |
| Very satisfied | 25 | 7.9 |
| **Describe your emotions/feelings during the PBF period** |  |  |
| Very unexcited | 18 | 5.7 |
| Unexcited | 52 | 16.5 |
| Neutral | 112 | 35.6 |
| Excited | 101 | 32.1 |
| Very excited | 32 | 10.2 |
| **How easy was it to implement and adapt to the PBF principles** |  |  |
| Very difficult | 23 | 7.3 |
| Difficult | 93 | 29.5 |
| Neutral | 71 | 22.5 |
| Easy | 111 | 35.2 |
| Very easy | 17 | 5.4 |
| **How appropriate was PBF for the health field** |  |  |
| Very inappropriate | 26 | 8.3 |
| Inappropriate | 39 | 12.4 |
| Neutral | 47 | 14.9 |
| Appropriate | 162 | 51.4 |
| Very appropriate | 41 | 13.0 |
| **How likely are you to recommend PBF in the future** |  |  |
| Very unlikely | 26 | 8.3 |
| Unlikely | 38 | 12.1 |
| Neutral | 35 | 11.1 |
| Likely | 142 | 45.1 |
| Very likely | 74 | 23.5 |

#### Error! Reference source not found.. Error! Reference source not found

Of the practitioners, 8.6% were very unsatisfied, 24.1% were just satisfied, 33.3% were neutral, 26% were satisfied, and 7.9% were very satisfied. The majority were neutral or just satisfied. Regarding emotions during the PBF period, 5.7% were very unexcited, 16.5% were unexcited, 35.6% were neutral, 32.1% were excited, and 10.2% were very excited, with most being neutral or excited. For ease of adaptability, 7.3% found it very difficult, 29.5% found it difficult, 22.5% were neutral, 35.2% found it easy, and 5.4% found it very easy, indicating it was generally easy to implement PBF. In terms of appropriateness, 8.3% found it very inappropriate, 12.4% inappropriate, 14.9% neutral, 51.4% appropriate, and 13% very appropriate. Regarding future recommendations, 8.3% were very unlikely to recommend PBF, 12.1% unlikely, 11.1% neutral, 45.1% likely, and 23.5% very likely, showing a positive inclination towards recommending PBF.

To calculate the Net Promoter Score (NPS) based on the provided survey responses, we need to categorize the scores as follows: Scores of 1 and 2 are considered "Distractors." A score of 3 is classified as "Passives." Scores of 4 and 5 are labeled as "Promoters."

The NPS for the question "How likely are you to recommend PBF in the future?"

Distractors: 26 (score 1) + 38 (score 2) = 64

Passives: 35 (score 3)

Promoters: 142 (score 4) + 74 (score 5) = 216

Calculate the percentage of Promoters by subtracting the percentage of Distractors from the percentage of Promoters:

NPS = (Promoters - Distractors) / Total Responses * 100.

NPS = (216–64) / (216 + 35 + 64) * 100 NPS = 152 / 315 * 100 NPS **≈ 48.25**

### Perception of Performance Based Financing

Assessing the entirety of results pertaining to the perception of health practitioners as per the PBF program (Table 3), the researcher realized that 62(19.7%) of the respondents perceived PBF as good, 169(53.7%) of the respondents perceived it as poor and 84(26.7%) of the respondents perceived it as very poor. With the modal score between 60-70%, one could conclude that; overall, the health practitioners perceived PBF as poor in enhancing output of health practitioners.

**Table 3:**
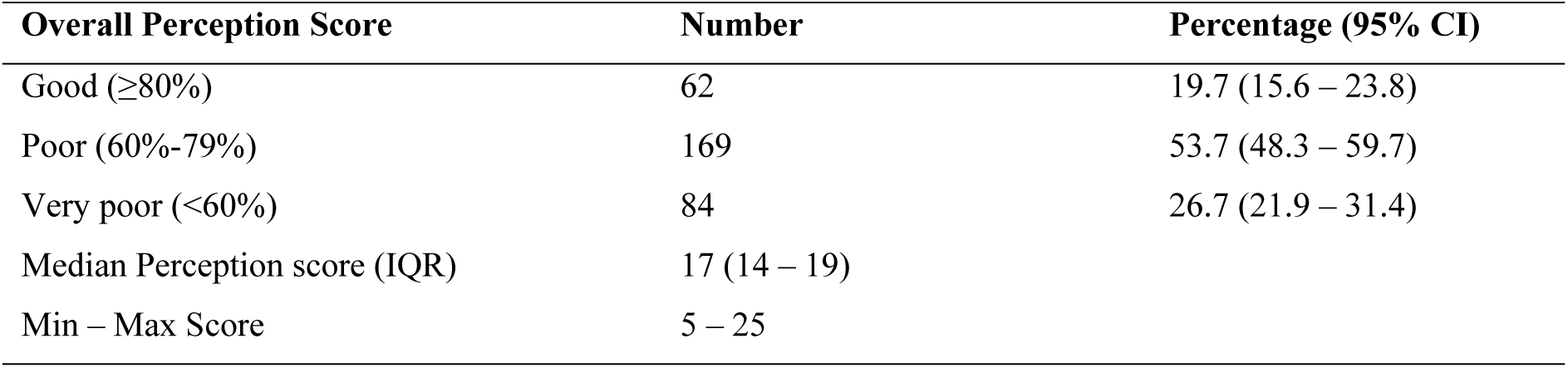
Perception Score.

| Overall Perception Score | Number | Percentage (95% CI) |
| --- | --- | --- |
| Good ($\geq 80\%$) | 62 | 19.7 (15.6 – 23.8) |
| Poor (60%-79%) | 169 | 53.7 (48.3 – 59.7) |
| Very poor (<60%) | 84 | 26.7 (21.9 – 31.4) |
| Median Perception score (IQR) | 17 (14 – 19) |  |
| Min – Max Score | 5 – 25 |  |

### Perceptions of Performance Based Financing based on various demographic and professional characteristics

The assessment of PBF perception by health practitioners revealed varied scores across districts and demographics (Table 4). In Bafut, 52% rated PBF very poor, while 52.6% in Bali and 58.3% in Bamenda scored it as poor. Nkwen had 45% rating it poor, Santa had 56.3%, and Tubah had 51%. Across age groups, those under 30 (69.6%) and those aged 30-39 (45.4%) predominantly scored PBF as poor. Gender-wise, 42.5% of males and 57.4% of females rated PBF as poor. Regarding education, 55.8% of university-educated practitioners and 63.9% of postgraduates scored PBF poorly. Functionally, 56.5% of nurses and 75% of doctors had negative scores. Overall, PBF received poor scores across districts, age groups, genders, education levels, and functions.

**Table 4:** Factors associated with perception.

| Variables | Perception score |  |  | P value |
| --- | --- | --- | --- | --- |
|  | Very poor<br>n (%) | Poor<br>n (%) | Good<br>n (%) |  |
| **Health District** |  |  |  | <0.001 |
| Bafut | 13 (52.0) | 11 (44.0) | 1 (4.0) |  |
| Bali | 2 (10.5) | 10 (52.6) | 7 (36.8) |  |
| Bamenda | 35 (26.5) | 77 (58.3) | 20 (15.2) |  |
| Nkwen | 18 (45.0) | 18 (45.0) | 4 (10.0) |  |
| Santa | 8 (16.7) | 27 (56.3) | 13 (27.1) |  |
| Tubah | 8 (15.7) | 26 (51.0) | 17 (33.3) |  |
| **Age groups** |  |  |  | 0.025 |
| <30 | 15 (19.0) | 55 (69.6) | 9 (11.4) |  |
| 30 – 39 | 40 (33.6) | 54 (45.4) | 25 (21.0) |  |
| 40 – 49 | 25 (28.1) | 43 (48.3) | 21 (23.6) |  |
| 50 – 59 | 4 (16.7) | 15 (62.5) | 5 (20.8) |  |
| $\geq 60$ | 0 (0.0) | 2 (50.0) | 2 (50.0) | |
| **Sex** |  |  |  | 0.067 |
| Male | 26 (32.5) | 34 (42.5) | 20 (25.0) |  |
| Female | 58 (24.7) | 135 (57.4) | 42 (17.9) |  |
| **Level of education** |  |  |  | 0.001 |
| Primary | 0 (0.0) | 5 (45.5) | 6 (54.5) |  |
| Secondary | 18 (40.9) | 14 (31.8) | 12 (27.3) |  |
| University | 52 (26.1) | 111 (55.8) | 36 (18.1) |  |
| Post graduate | 14 (23.0) | 39 (63.9) | 8 (13.1) |  |
| **Function** |  |  |  | <0.001 |
| Nurse | 43 (22.3) | 109 (56.5) | 41 (21.2) |  |
| Lab Tech | 9 (34.6) | 7 (26.9) | 10 (38.5) |  |
| Midwife | 15 (41.7) | 21 (58.3) | 0 (0.0) |  |
| Biomed tech/Imaging | 1 (33.3) | 2 (66.7) | 0 (0.0) |  |
| Medical doctor | 6 (75.0) | 2 (25.0) | 0 (0.0) |  |
| **Auxiliary** | 10 (20.4) | 28 (57.1) | 11 (22.4) |  |

### The effect of PBF on health workers’ output

The data on Table 5 shows significant shifts in how respondents rated their output before and during the PBF implementation across key variables. For general workload, high and very high ratings increased from 26% before PBF to 66.9% during PBF, while ratings for very low, low, and acceptable workloads dropped from 73.9% to 33%. High and very high ratings for monthly patient consultations surged from 43.8% before PBF to 78.1% during PBF, with low and acceptable consultations falling from 56.2% to 21.9%. Monthly admissions rated high and very high rose from 33% before PBF to 63.5% during PBF, with lower ratings decreasing from 67% to 36.5%. For monthly deliveries, high and very high ratings increased from 32.7% before PBF to 61.6% during PBF, while lower ratings fell from 67.3% to 38.4%. Community interventions saw high and very high ratings rise from 21.6% before PBF to 50.5% during PBF, with lower ratings declining from 78.4% to 49.5%. High and very high financial incentives increased from 9.8% before PBF to 33% during PBF, while lower incentives dropped from 90.2% to 67%. Overtime ratings for high and very high levels grew from 32.4% before PBF to 59.7% during PBF, with lower overtime ratings decreasing from 67.6% to 40.3%. No significant change was noted in monthly mortality rates (p=0.055), although very low mortality rates were slightly higher during PBF (38.4%) compared to before PBF (34.6%). Overall, respondents indicated increased activity and output during the PBF period, with higher ratings for workloads, consultations, admissions, deliveries, community interventions, financial incentives, and overtime.

**Table 5:** Effect of Performance Based Financing on Output.

| **Variables** | **Before PBF n (%)** | **During PBF n (%)** | **P value** |
| --- | --- | --- | --- |
| **General workload** |  |  | <0.001 |
| Very low | 22 (7.0) | 2 (0.6) |  |
| Low | 54 (17.1) | 11 (3.5) |  |
| Acceptable | 157 (49.8) | 91 (28.9) |  |
| High | 65 (20.6) | 157 (49.8) |  |
| Very High | 17 (5.4) | 54 (17.1) |  |
| **Average number of patients consulted monthly** |  |  | <0.001 |
| Very low | 5 (1.6) | 0 (0.0) |  |
| Low | 42 (13.3) | 2 (0.6) |  |
| Acceptable | 130 (41.3) | 67 (21.3) |  |
| High | 111 (35.2) | 168 (53.3) |  |
| Very High | 27 (8.6) | 78 (24.8) |  |
| **Average number of patients admitted monthly** |  |  | <0.001 |
| Very low | 12 (3.8) | 0 (0.0) |  |
| Low | 43 (13.7) | 4 (1.3) |  |
| Acceptable | 156 (49.5) | 111 (35.2) |  |
| High | 88 (27.9) | 118 (37.5) |  |
| Very High | 16 (5.1) | 82 (26.0) |  |
| **Average number of monthly deliveries** |  |  | <0.001 |
| Very low | 13 (4.1) | 5 (1.6) |  |
| Low | 63 (20.0) | 14 (4.4) |  |
| Acceptable | 136 (43.2) | 102 (32.4) |  |
| High | 84 (26.7) | 131 (41.6) |  |
| Very High | 19 (6.0) | 63 (20.0) |  |
| **Number of monthly community interventions** |  |  | <0.001 |
| Very low | 13 (4.1) | 1 (0.3) |  |
| Low | 104 (33.0) | 36 (11.4) |  |
| Acceptable | 130 (41.3) | 119 (37.8) |  |
| High | 51 (16.2) | 112 (35.6) |  |
| Very High | 17 (5.4) | 47 (14.9) |  |
| **Average monthly financial incentives** |  |  | <0.001 |
| Very low | 66 (21.0) | 38 (12.1) |  |
| Low | 128 (40.6) | 62 (19.7) |  |
| Acceptable | 90 (28.6) | 111 (35.2) |  |
| High | 23 (7.3) | 80 (25.4) |  |
| Very High | 8 (2.5) | 24 (7.6) |  |
| **Monthly mortality rate** |  |  | 0.055 |
| **Very low** | 109 (34.6) | 121 (38.4) |  |
| **Low** | 102 (32.4) | 90 (28.6) |  |
| **Acceptable** | 63 (20.0) | 51 (16.2) |  |
| **High** | 33 (10.5) | 31 (9.8) |  |
| **Very High** | 8 (2.5) | 22 (7.0) |  |
| **Overtime** |  |  | <0.001 |
| **Very low** | 29 (9.2) | 14 (4.4) |  |
| **Low** | 61 (19.4) | 34 (10.8) |  |
| **Acceptable** | 123 (39.0) | 79 (25.1) |  |
| **High** | 61 (19.4) | 109 (34.6) |  |
| **Very High** | 41 (13.0) | 79 (25.1) |  |

### The effects of Performance Based Financing on healthcare quality

The PBF project significantly positively impacted various quality indicators as rated by health practitioners(Table 6) For patient waiting times, the number of respondents reporting wait times under 30 minutes increased from 35.6% before PBF to 55.9% during PBF, while those reporting over 60 minutes decreased from 25.7% to 14%. Hospital cleanliness improved markedly, with ratings of ’very clean’ increasing from 12.1% before PBF to 50.2% during PBF and ’very dirty’ dropping from 16.2% to 0.3%. Equity of care saw substantial improvements, with high and very high ratings rising from 38.7% before PBF to 73.4% during PBF. Assiduity ratings also improved, with high and very high levels increasing from 30.8% to 74% during PBF. Patient reception quality saw a similar trend, with high and very high ratings rising from 31.4% to 70.8%. Self-efficacy among health practitioners improved, with high and very high levels increasing from 38.5% to 76.5%. Infrastructural development ratings improved significantly, with high and very high levels rising from 34.3% to 77.1% during PBF. Patient extortion and embezzlement ratings showed a notable decrease, with high and very high levels dropping from 25.7% to 9.5%. Overall, the PBF project led to considerable enhancements in the quality of healthcare delivery as perceived by health practitioners.

**Table 6:** Effect of Performance based Financing on Healthcare.

| **Variables** | **Before PBF n (%)** | **During PBF n (%)** | **P value** |
| --- | --- | --- | --- |
| **Approximate patient waiting time** |  |  | <0.001 |
| Less than 30 mins | 112 (35.6) | 176 (55.9) |  |
| 30 – 60 mins | 122 (38.7) | 95 (30.2) |  |
| Greater than 60 mins | 81 (25.7) | 44 (14.0) |  |
| **Cleanliness of the hospital** |  |  | <0.0001 |
| Very dirty | 51 (16.2) | 1 (0.3) |  |
| Dirty | 63 (20.0) | 7 (2.2) |  |
| Acceptable | 81 (25.7) | 33 (10.5) |  |
| Clean | 82 (26.0) | 116 (36.8) |  |
| Very clean | 38 (12.1) | 158 (50.2) |  |
| **Equity of care** |  |  | <0.001 |
| Very low | 16 (5.1) | 0 (0.0) |  |
| Low | 46 (14.6) | 4 (1.3) |  |
| Acceptable | 131 (41.6) | 80 (25.4) |  |
| High | 69 (21.9) | 136 (43.2) |  |
| Very high | 53 (16.8) | 95 (30.2) |  |
| **Assiduity** |  |  | <0.001 |
| **Very low** | 23 (7.3) | 0 (0.0) |  |
| **Low** | 51 (16.2) | 10 (3.2) |  |
| **Acceptable** | 144 (45.7) | 72 (22.9) |  |
| **High** | 74 (23.5) | 144 (45.7) |  |
| **Very high** | 23 (7.3) | 89 (28.3) |  |
| **Quality of patient reception** |  |  | <0.001 |
| **Very low** | 18 (5.7) | 10 (3.2) |  |
| **Low** | 61 (19.4) | 2 (0.6) |  |
| Acceptable | 137 (43.5) | 80 (25.4) |  |
| High | 76 (24.1) | 139 (44.1) |  |
| Very high | 23 (7.3) | 84 (26.7) |  |
| **Self-efficacy (work confidence)** |  |  | <0.001 |
| Very low | 14 (4.4) | 2 (0.6) |  |
| Low | 46 (14.6) | 10 (3.2) |  |
| Acceptable | 135 (42.9) | 62 (19.7) |  |
| High | 76 (24.1) | 142 (45.1) |  |
| Very high | 44 (14.0) | 99 (31.4) |  |
| **Infrastructural development** |  |  | <0.001 |
| Very low | 25 (7.9) | 13 (4.1) |  |
| Low | 63 (20.0) | 15 (4.8) |  |
| Acceptable | 119 (37.8) | 44 (14.0) |  |
| High | 44 (14.0) | 134 (42.5) |  |
| Very high | 64 (20.3) | 109 (34.6) |  |
| **Patient extortion and embezzlement** |  |  | <0.001 |
| Very low | 108 (34.3) | 165 (52.4) |  |
| Low | 79 (25.1) | 83 (26.3) |  |
| Acceptable | 47 (14.9) | 37 (11.7) |  |
| High | 60 (19.0) | 20 (6.3) |  |
| Very high | 21 (6.7) | 10 (3.2) |  |

Source: Researcher, 2023

### Effect of Performance based Financing on Healthcare Quality

Scoring the quality of the results obtained as a result of the PBF project, the results were grouped and scored under the brackets of good (80%), poor (60%-79%), and very poor (<60%). Table 7 shows that Good quality ratings increased from 17.8% before PBF to 59.4% during PBF. Poor quality scores dropped from 41.9% to 36.2%, and very poor ratings decreased from 40.3% to 4.4%. This demonstrates that the PBF project significantly improved service quality in the health districts.

**Table 7:** Overall healthcare quality score for periods before and during the implementation of the Performance Based Financing Project.

| Quality score | Before PBF<br>n (%) | During PBF<br>n (%) | P value |
| --- | --- | --- | --- |
| Good ($\geq 80\%$) | 56 (17.8) | 187 (59.4) | <0.001 |
| Poor (60% - 79%) | 132 (41.9) | 114 (36.2) |  |
| Very poor (< 60%) | 127 (40.3) | 14 (4.4) |  |
| **Median Quality score (IQR)** | 25 (21 – 29) | 32 (28 – 34) | <0.001 |

Source: Researcher, 2023

### Summary of findings

This study examines the impact of PBF on healthcare workers in the Mezam Health District of Cameroon, revealing significant demographic and professional insights. Predominantly female, the surveyed group had a substantial number holding university degrees, including nurses, lab techs, midwives, and doctors. Perceptions of PBF varied by district, age, and role, with nurses and lab techs viewing it more positively although many view PBF as a poor strategy, PBF led to increased workloads, more patients consulted, admitted, and delivered, and expanded community interventions. Financial incentives improved, albeit with increased overtime. Importantly, PBF enhanced healthcare quality, reducing patient waiting times and improving hospital cleanliness, equity of care, assiduity, patient reception quality, self-efficacy, and infrastructural development while significantly decreasing patient extortion and embezzlement. Overall, the study underscores PBF’s potential to boost healthcare worker performance and service quality despite increased workloads.

## DISCUSSION OF FINDINGS

### Socio-Demographics

The study identified significant demographic variations among healthcare workers in Mezam across districts, age groups, and gender. Bamenda had the highest number of healthcare workers (41.9%), while Bafut had the lowest (7.9%), reflecting proportional sampling, as was done in Ghana (40).

Most healthcare workers were aged 30-39 years (37.8%), consistent with trends in sub-Saharan Africa (41). Females made up the majority of the workforce (74.6%), aligning with the global trend of female dominance in nursing and midwifery (1), likely due to a natural inclination towards caregiving roles. The majority of healthcare workers had university degrees (63.2%), with 19.4% holding postgraduate qualifications, reflecting the high educational requirements of the field. This distribution is consistent with findings from LMICs, such as Ethiopia (42). Among healthcare functions, nurses were the most prevalent (61.3%), followed by Auxiliaries (15.6%) and Midwives (11.4%), aligning with global trends where nursing constitutes a significant portion of the healthcare workforce (43).

During PBF, the distribution of healthcare workers by employer type showed minor changes, with a slight increase in government employment and decreases in the private sector and confessional organizations, though these shifts were not statistically significant (p = 0.062). Similar studies in low- and middle-income countries (LMICs) have also noted changes in employment patterns following health system reforms (44). Conversely, the distribution by healthcare facility type changed significantly, with more workers in district and regional hospitals and fewer in health centers (p = 0.019), reflecting trends observed in LMICs where workers favor better-equipped facilities (45). Marital status remained stable, with a slight increase in married workers (p = 0.607), potentially due to job stability (46). The religious composition shifted significantly towards more Christian workers (p = 0.046), while longevity and income also increased, correlating with job satisfaction and financial incentives (47); (48). Household sizes grew significantly p < 0.001, possibly due to factors like time and displacement due to the sociopolitical crisis (49)

### Healthcare workers’ perceptions and attitudes toward the Performance Based Financing project

Healthcare workers expressed mixed feelings regarding whether their expectations from Performance-Based Financing (PBF) were met, with 33.3% remaining neutral and others showing varying satisfaction levels. This aligns with findings from other PBF programs, indicating diverse expectations and experiences among healthcare workers (50). During the PBF period, emotional responses varied: 35.6% felt "Neutral" while 32.1% were "Excited," reflecting the complexity of PBF implementation, where some embraced changes and financial incentives while others faced uncertainties (13). The importance of leadership and organizational capacity in shaping health workers’ reactions to PBF is vital (34).

Opinions on the ease of implementing and adapting to PBF principles were also mixed; 35.2% found it "Easy" while 29.5% found it "Difficult." This highlights the need for effective training and support to help healthcare workers transition smoothly (22). Regarding PBF’s appropriateness for healthcare, 51.4% deemed it "Appropriate," but 12.4% viewed it as "Inappropriate," suggesting that contextual factors influence perceptions (50). Similar studies, such as those by (22)in Rwanda, showed more positive views, highlighting the importance of context. In terms of future recommendations, 45.1% were "Likely" to recommend PBF, while 12.1% were "Unlikely," suggesting both potential endorsement and concerns that need addressing for PBF’s broader acceptance (50).

The calculated Net Promoter Score (NPS) of approximately 48.25% suggests a positive perception of PBF among healthcare workers, with more promoters than detractors. This positive NPS suggests that, based on the responses to the question about perception of PBF, there is a higher percentage of promoters than detractors, indicating that healthcare workers generally have a positive perception of PBF. The positive perception of PBF among healthcare workers have been noted in various settings (51). Moreover, the usability and acceptance of PBF among health workers can be improved by ensuring timely and adequate payouts of performance-based payments (52).

### Perceptions of Performance Based Financing based on various demographic and professional characteristics

This study examined healthcare workers’ perceptions of PBF across different demographic and professional characteristics, focusing on health district, age, sex, education level, and occupation. Perceptions varied significantly by health district, possibly due to differences in PBF implementation strategies, stakeholder engagement, cultural norms, and socio-economic conditions. Health facilities with stronger systems viewed PBF positively, while weaker ones saw it as burdensome, aligning with findings by (53) and (34)

Younger healthcare workers tended to be more critical of PBF, potentially due to their technological proficiency and expectations for participation and innovation. In contrast, adults had mixed opinions, influenced by their experience with similar systems and less technological familiarity (54); (41). Gender differences also emerged: males had more negative perceptions, citing concerns about accountability, equity, and job security, while females viewed PBF more positively, valuing competition, rewards, and gender equality (55); (56); (41).

Education level influenced perceptions, with more educated healthcare workers showing positive views due to a better understanding of funding challenges and alignment with organizational goals (57). However, contrary to (42)doctors generally had negative perceptions, possibly due to concerns about quality care, administrative burden, and misalignment of goals, while nurses and lower-level staff were more positive.

Overall, the study noted significant variations in perceptions across demographic groups, similar to findings by (22)in Rwanda, where healthcare workers generally viewed PBF positively, and (50)who found mixed responses in a multi-country study. While PBF can motivate health workers to improve care quality, certain barriers beyond their control can hinder the effectiveness of these programs (58). The relationship between financial incentives and health worker motivation is complex, as evidenced by the need to consider various factors influencing workers’ reactions to PBF initiatives (59)

### The effect of Performance Based Financing on health workers’ output

General workload increased under PBF, with more health workers reporting a "High" workload compared to the "Acceptable" levels before PBF (p < 0.001). This finding contrasts with some literature suggesting that PBF can enhance resource allocation efficiency (60), likely due to local contextual factors. The average number of patients consulted monthly also rose significantly, with a shift from "Acceptable" to "High" patient loads (p < 0.001), aligning with studies in Tanzania and Zambia that found increased service delivery due to PBF (24); (61)

Similarly, monthly admissions and deliveries saw significant increases (p < 0.001), driven by PBF’s focus on equity and promoting facility-based care. Community interventions and financial incentives also improved, indicating that PBF incentivizes engagement and rewards performance (22); (62); (63). However, the monthly mortality rate showed no significant change (p = 0.055), suggesting that increased service delivery did not immediately translate into improved mortality. However, there could be a reduction in community death.

PBF significantly increased overtime levels, with many health workers reporting "High" overtime (p < 0.001), which raises concerns about the potential negative impact on their well-being and work-life balance (24). These results echo findings from other studies, showing that while PBF can enhance health worker output and service quality, its effectiveness varies across different settings and is influenced by contextual factors, financial incentives, and facility capacity/application (25); (23); (64)

The implementation of PBF in Mezam has had a significant positive impact on various aspects of healthcare quality. Notably, PBF led to a substantial reduction in patient waiting times, with 55.9% of patients experiencing waits of less than 30 minutes during PBF compared to 35.6% before (p < 0.001). Hospital cleanliness also improved, with more respondents perceiving facilities as "Clean" or "Very Clean" during PBF (p < 0.0001), contrasting with some literature suggesting inefficient use of PBF funds for infrastructure (62); (21); (32); (33)

PBF also positively impacted the perceived equity of care, with a significant increase in ratings of "High" equity (p < 0.001), supporting findings from (65). The assiduity, or diligence, of healthcare workers improved, with higher perceptions of diligence during PBF (p < 0.001), aligning with studies by (22). Furthermore, the quality of patient reception and healthcare workers’ self-efficacy saw significant boosts during PBF, reflecting increased patient satisfaction and worker confidence (p < 0.001).

Infrastructure development perceptions also improved significantly during PBF (p < 0.001), suggesting effective use of funds contrary to some previous findings (62). Moreover, PBF was associated with a marked reduction in patient extortion and embezzlement, with 52.4% reporting "Very Low" occurrences during PBF (p < 0.001), aligning with studies highlighting PBF’s role in reducing corruption ((60); (66). These findings reflect the potential of PBF to enhance healthcare service quality and efficiency, echoing similar outcomes observed in Rwanda and other settings (22); (14). However, contextual factors and implementation challenges must be considered, as mixed results have been reported in other studies (12)

### Validation of Hypotheses

Hypothesis 1, suggesting no difference in health workers’ perceptions based on demographics, was rejected. Findings showed significant variations in perceptions influenced by health district, age, and education level, with more educated workers viewing PBF more positively. We therefore reject the null hypothesis and conclude that there are significant variations in perceptions of Performance Based Financing.

Hypothesis 2, proposing no relationship between PBF implementation and health workers’ output, was also rejected. Significant differences in workload, patient load, admissions, and financial incentives between "Before PBF" and "During PBF" periods indicated a clear impact of PBF on output. Therefore, we rejected the null hypothesis and concluded that there were significant differences in workload, patient load, admissions, and finances before and during the performance-based financing project’s implementation.

Hypothesis 3, which posited no difference in healthcare quality before and after PBF. Statistically significant improvements were observed in patient waiting times, cleanliness, healthcare equity, and reductions in patient extortion and embezzlement, demonstrating that PBF positively affected healthcare quality. The null hypothesis was thus rejected and the study concludes that there are statistically significant differences in healthcare quality before and During the Performance Based funding implementation process.

## CONCLUSION AND RECOMMENDATIONS

### Conclusion

The study reveals that health workers in Mezam have a generally positive perception of the PBF project, viewing it as a beneficial initiative that enhances their motivation, job satisfaction, and performance, largely due to the financial incentives provided. These incentives play a crucial role in boosting productivity, as evidenced by increased patient consultations, admissions, deliveries, and community interventions. Moreover, the PBF project positively impacts healthcare quality, with health workers reporting greater adherence to clinical protocols and improved care delivery. The emphasis on quality assurance and regular monitoring within the PBF framework further supports these positive outcomes.

### Recommendations

Several key recommendations can be made Based on the study’s findings on the impact of the PBF project in Mezam Health District. First, sustaining and expanding the PBF project is essential, as it has positively influenced healthcare worker output and service quality; ongoing adjustments based on performance metrics and feedback should be implemented to enhance its effectiveness further. Second, optimizing workload management is crucial to address the increased demands associated with PBF; strategies such as hiring more staff, improving scheduling systems, and providing training can help ensure efficient workload distribution and prevent burnout. Third, enhancing financial incentives is necessary, as the increase in overtime suggests a need to review the incentive structure to motivate workers without leading to excessive hours, thereby promoting a healthy work-life balance. Fourth, continuing to focus on quality and equity is vital; efforts to improve cleanliness, patient reception, and equitable care should be maintained alongside robust mechanisms to monitor and address any issues related to patient extortion and embezzlement. Fifth, integrating the PBF model into the Universal Health Coverage (UHC) policy presents an opportunity to leverage its success in improving healthcare quality and worker performance, supporting the expansion of UHC in Cameroon through targeted financial incentives that enhance both access to care and service quality. The study demonstrates that improvements in equity of care and system efficiency under PBF can guide policymakers in designing Universal Health Coverage strategies that address healthcare disparities, optimize resource use, and enhance trust in healthcare services through transparent financial systems.

## Data Availability

Data is attached herein. Further information can be gotten from the authors via their emails.

## REFERENCES

1. World Health Organisation. world Health Statistics 2021: A Visual summary. 2021.

2. world Health Organisation. world Health statistics 2017: monitoring health for the SDGs. 2017.

3. Trends in Health expenditure in the United states and Canada from 1996 to 2013: A tale of 2 countries. Lu, C., et al. 6, 2021, Health Affairs, Vol. 30, pp. 872–882.

4. World Health Organisation;. *Universal Health Coverage*. 2019.

5. Performance-based financing in low and middle-income countries: A systematic review of its effectiveness in improving health outcomes. Kellett, S. 11, s.l. : 10.34172/ijhpm.2020.189, 2020, International Journal of Health Policy and Management, Vol. 9, pp. 462–473.

6. Performance-Based Financing: A review of the evidence and implications for health systems. DFC. 2021, DFC research paper, Vol. 2021/001.

7. Performance Based incentives for health: A way to improve tuberculosis detection and treatment completion. Cashin, C., Sparkes, S. and Bloom, D. 1, 2014, Health systems and Reforms, Vol. 1, pp. 66–74.

8. Understanding Private general practitioners’ perspectives on China’s health reform. Hood, C. M., Goudge, J. and Su, Y. 1, 2018, International Journal for Equity in Health, Vol. 17, pp. 1–11.

9. Savedoff, W. D. and Hussmann, K. *Why are health systems prone to corruption?* Education. s.l. : Global corruption report, 2020.

10. The importance of leadership and organizational capacity in shaping health workers’ motivational reactions to performance-based financing: a multiple case study in burkina faso. Fillol, A., et al. 5, s.l. : 10.15171/ijhmp.2018.133, 2019, International Journal of Health Policy and Management, Vol. 8, pp. 272–279.

11. Healthcare digitalization and pay-for-performance incentives in smart hospital project financing. Visconti, R. and Morea, D. 7, s.l. : 10.3390/ijerph17072318, 2020, International Journal of Environmental Research and Publiv Health, Vol. 17, p. 2318.

12. Context matters(but how and why?) A hypothesis-led literature review of performance based financing in fragile and conflict-affected health systems. P. Bertone, M. P., et al. 4, 2018, PLoS ONE, Vol. 13. e0195301.

13. Performance-based financing for better quality services in Rwandan health centres: 3-years Experience. Renmans, D., Holvoet N. and Orach, C. G. 4, 2017, Tropical Medicine & International Health, Vol. 22, pp. 348–356.

14. Witter, S., et al. Paying for performance to mprove the delivery of health interventions in low and middle-income countries. Cochrane datavase of systemetic reviews. 2012, p. 2.

15. Fritche, G. B., Soeters, R. and Meessen, B. Performance Based financing toolkit. [Online] 2014. https://openknowledge.worldbank.org/handle/10986/17194.

16. Performance Based Financing: A Catalyst for improving Healthcare Services in Cameroon. Njikam, O., Fofung, E. and Fomba, M. J. 2, 2021, Health Systems and Reforms, Vol. 7, pp. 115–120.

17. Ministry of Public Health Cameroon. *National Health development plan(2022-2026)*. Ministry of Public Health. 2022. https://www.minsante.cm/site/?q=en/content/national-health-development-plan-2022-2026.

18. MOtivating Health workers up to a limit: partia; effects of performance-based financing on working environments in Nigeria. Bhatnagar, A. and George, A. 7, s.l. : 10.1093/heapol/czw002, 2016, Health Policy and Planning, Vol. 31, pp. 868–877.

19. Aninanya, G., et al. Can performance-based incentives improve motivation of nurses and midwives in primary facilities in northern ghana? a quasi-experimental study. Global Health Action. 2016, Vol. 9, 1.

20. Implementation Research to improve quality of maternal and newborn health care, malawi. Brenner, S., et al. 7, s.l. : 10.2471/blt.16.178202, 2017, Bulletin of the orld Health Organisation, Vol. 95, pp. 491–502.

21. Performance Based Financing to strengthen the Health System in Benin: Challenging the mainstream approach. Paul, E., et al. 1, s.l. : 10.15171/ijhpm.2017.42, 2017, International Journal of Health Policy and Management, Vol. 7, pp. 35–47.

22. Effect on Maternal child health services in Rwanda of payment to primary health-care providers for performance: an impact evaluation. Basinga, P., et al. 9775, s.l. : 10.1016/s0140-6736(11)60177-3, 2011, The Lancet, Vol. 377, pp. 1412–1428.

23. Using targeted vouchers and health equity funds to improve access to skilled birth attendants for poor women: A case study in three rural health districts in Cambodia. Sabri, B., et al. 1, s.l. : 10.1186/1471-2393-10-1, 2007, BMC Pregnancy and Childbirth, Vol. 7.

24. Using performance incentives to improve medical care productivity and health outcomes. Gertler, P. J., Vermeersch, C. M. and Van, Der Gaag. 2015, National Bureau of Economic Research.

25. Reviewing institutions of rural Health centers: the performance initiative in Butare, Rwanda. Meessen, B., et al. 8, 2006, Tropical Medicine & international Health, Vol. 11, pp. 1303–1317. 1303-1317. doi:10.1111/j.1365-3156.2006.01680.x..

26. HIV treatment in the private sector: An analysis of ART service delivery in Zambia. Friedman, J., et al. 1, 2016, AIDS Research Therapy, Vol. 13, p. 25.

27. United Nations. Transforming our world: the 2030 agenda for Sustainable Development,. New York:UN : s.n., 2015.

28. Towards a coherent global framework for health financing: recommendations and recent developments. Ottersen, T., et al. 2, s.l. : 10.1017/S1744133116000505., 2017, Health Economics policy law, Vol. 12, pp. 285–296.

29. Setting performance-based financing in the health sector agenda: a case study in Cameroon. Globalization and Health. Sieleunou, I., et al. 2017, p. 13.

30. Incentices to change:Effecs of performance-based financing on health in Zambia. Shen, G., et al. 1, s.l. : 10.1186/s12960-017-0179-2, 2017, Human Resources for Health, Vol. 15.

31. Performance-Based financing kick starts motivational "feedback loop": findings from a process evaluation in Mozambique. Gergen, J., et al. 1, s.l. : 10.1186/s12960-018-0320-x, 2018, Human Resources for Health, Vol. 16.

32. Can Performance based financing be used to reform health systems in developing countries? Ireland, M., Paul, E. and Dujardin, B. 9, s.l. : 10.2471/blt.11.087379, 2011, Bulletin of the world health organisation, Vol. 89, pp. 695–698.

33. Does training on performance based financing make a difference in performance and quality of health care delivery? health care provider’s perspective in rungwe Tanzania. Manongi, R., et al. 1, s.l. : 10.1186/1472-6963-14-154, 2014, BNC Health Services, Vol. 14.

34. The importance of leadership and organisational capacity in shaping health workers’ motivational reactions to performance-based financing: a multiple case study in Burkinafase. Fillol, A., et al. 5, s.l. : 10.15171/ijhpm.2018.133, 2019, International Journal of Health Policy and Management, Vol. 8, pp. 272–279.

35. *Performance-based financing for healthcare in the Northwest Region, Cameroon: A population-based study*. CIS. s.l. : 10.131140/RG.2.2.23558.93124, 2022, CIS report 2022–2.

36. RDPH. *Health system Performance in Northwest Region, Cameroon: A review of Progress Towards Universal Health Coverage*. NWR : RDPH, 2023.

37. Socio-political crisis and healthcare access in Northwest Region, Cameroon: A qualitative study. Bang, H. T. and Balgah, R. Z. 2, s.l. : 10.1177/00207313221082942, 2022, International Journal of Health, Vol. 52, pp. 237–248.

38. Performance-Based Financing Toolkit. World Bank. s.l. : http://documents1.worldbank.org/curated/en/358251467994882004/pdf/940550v10WP0Box385358B00PUBLIC0.pdf, 2015, World Bank.

39. INS. Health Insurance Coverage in Cameroon:A Descriptive Analysis. s.l. : INS report 2019–02, 2019.

40. Health worker (internal customer) satisfaction and motivation in the public sector in Ghana. Agyepong, L. A., et al. 2, s.l. : 10.1002/hpm.770, 2012, International Journal of Health Planning and Mangement, Vol. 27, pp. 163–177. https://doi.org/10.1093/hpm.770.

41. English language proficiency as a barrier to understanding health information and services in Kenya: A study of the Kamba people of Kitui District. Mbindyo, P., Blaauw, D. and Gilson, L. 5, 2009, Health Policy and Planning, Vol. 24, pp. 418–428.

42. Physician distribution and arreition in the public health sector of Ethopia. Assefa, T., et al. s.l. : 10.2147/rmhp.s117943, 2017, Risk Management and Healthcare Policy, Vol. 10, pp. 17–29.

43. Impact and determinants of nurse turnover: A pan-Canadian study. O’Brien-Pallas, L., et al. 6, 2003, Journal of Nursing Management, Vol. 11, pp. 474–486.

44. The deterioration of formal employment opportunities in the wake of the economic crisis in Zambia: A multi-level analysis of the determinants of health workers’ choices. Sasaki, S., et al. 4, 2018, Health Policy and Planning, Vol. 33, pp. 505–513.

45. Operational readiness of public hospitals in northern Bangladesh to receive internally displaced persons: A cross-sectional study. Hossain, S. M. M., et al. 1, 2019, Conflict and Health, Vol. 13, p. 33.

46. Poverty and Health care: A review of the literature. GLobal Public Health,. Cohen, J. 2, s.l. : 10.1080/17441690500104042, 2005, Global Public Health, Vol. 1.

47. *Getting health workers to rural areas: Innovative analytical approaches to inform policy implementation*. Vujicic, M., Alfano, M. and Shengelai, B. washington DC : World Bank, 2011.

48. Population health and economic growth. Barnighausen, T., et al. 7, s.l. : 10.1016/j.worlddev.2003.07.002, 2008, World Development, Vol. 36, pp. 987–1016.

49. Health-care needs of people affected by conflict: Future trends and changing frameworks. Spiegel, P. B., et al. 9711, 2014, The Lancet, Vol. 375, pp. 341–345.

50. Assessing perceived quality of care as a predictor of health facility performance: A cross-sectional study of mid-level providers in Mozambique. Bertone, M. P., et al. 6, s.l. : 10.1371/journal.pone.0195301, 2016, PLoS ONE, Vol. 11. e0155207.

51. Manga, L., et al. Performance based financing and Job Satisfaction in a Semi-urban health district in Cameroon. Journal of Public Health in Africa. s.l. : 10.4081/jphia.2018.760, 2018.

52. Usability and acceptance of a mobile health wallet for pregnancy-related healthcare: a mixed methods study on stakeholders’ perceptions in central madagascar. Lacroze, E., et al. 1, s.l. : 10.1371/journal.pone.0279880, 2023, PLoS ONE, Vol. 18. e0279880.

53. A mixed method exploration of health workers attitudes towards Performance Based Funding in Burkina Faso. Lohmann, Julia, et al. 8, s.l. : PubMed Central, 2020, international Journal of health and policy Management, Vol. 10, pp. 483–494. 32610757.

54. Person-Organisation Fit and the War for Talent: Does it Matter? Ng, E. S. W. and Burke, R. J. 7, 2005, International journal of human resource Management, Vol. 16, pp. 1195–1210.

55. The Leadership Styles of Men and Women. Eagly, A. H. and Johannesen-Schmidt, M. C. 4, 2001, Journal of Social Issues, Vol. 57, pp. 781–797.

56. Work-Family Conflict, Policies, and job-life satisfaction Relationship: A review and Directions for future research. Kossek, E. E. and Ozeki, C. 2, 1998, Journal of Applied Psychology, Vol. 83, pp. 139–149.

57. Evaluating the quality of medical care. Donabedian, A. 3, 1966, The Milbank Memorial Fund Quaterly, Vol. 44, pp. 166–206.

58. Experiences of care in the context of payment for performance (P4P) in tanzania. Chimutu, V., Tjomsland, M. and Mrisho, M. 1, s.l. : 10.1186/s12992-019-0503-9, 2019, Globalisation and Health, Vol. 15.

59. Zitti, T., et al. Does the gap between healthh workers’ expectations and the realities of implementing a performance-based financing project in Mali create frustration? s.l. : 10.21203/rs.3.rs-54170/v3, 2021.

60. Reviewing institutions of rural health centres: The Performance Initiative in Butare, Rwanda. Meessen, B., et al. 11, 2011, Tropical Medicine and International Health, Vol. 16, pp. 1439–1446.

61. Analysis of Strategies to attract and retain rural health workers in Cambodia, China and Vietnam and context influencing their outcomes. Zhu, A., et al. 1, s.l. : 10.1186/s12960-018-0340-6, 2019, Human Resources for Health, Vol. 17.

62. The effects of performance incentives on the utilization and quality of maternal and child care in Burundi. Bonfrer, I., Van de Poel, E. and Van Doorslaer, E. 2014, Social Science & Medicine, Vol. 123, pp. 96–104.

63. Effects of comunity performance-based financing on comunity health workers’ service delivery in Kayanza health district, Burundi. Gervais, M., Odongo, A. and Maima, A. 1, s.l. : 10.18203/2394-6040.ijcmph20223521, 2022, International Journal of Comunity Medicine and Public Health, Vol. 10, p. 21.

64. Effects of performance-based financing on maternal care in 14 districts in Burundi. Soeters, R., Habineza, C. and Peerenboom, P. B. 2, 2006, The international Joutnal of Health Planning and Management, Vol. 26, pp. 214–226.

65. Why performance-based financing is saving lives in Rwanda. Sengooba, F., McPake, B. and Palmer, N. 3, s.l. : 10.1093/inthealth/ihw024, 3026, International Journal of health, Vol. 8, pp. 161–166.

66. Performance-based financing for better quality of services in Rwandan health centres: 3-year experience. Rusa, L., et al. 7, s.l. : doi:10.1111/j.1365-3156.2009.02292.x., 2009, Tropical Medicine and international Health TM IH, Vol. 14, pp. 830–837.

67. The Quality of Care: How can it be Assessed? Donabedian, A. 12, 1988, Journal of American Medical Association, Vol. 260, pp. 1743–1748.

68. incentives for quality improvement: the role of education in shaping perceptions of performance-based financing. Austin, A. E. and Bowers, A. A. 2016, BMC Health Services Research, Vol. 16, p. 543.

69. Transforming our world: The 2030 Agenda for Sustainable development. United Nations. United Nations : s.n., 2018.

70. *Sources and focus of health development assistance*, *1990-2014*. Dieleman, J. L. 23, 2016, JAMA, Vol. 313, pp. 2359–2368.

